# Cost-effectiveness of CTC guided chemo- or endocrine therapy in ER+ HER2- metastatic breast cancer – results from a randomized controlled multicenter trial

**DOI:** 10.1101/2023.10.09.23296711

**Authors:** A.M.S. Berghuis, H. Koffijberg, X.G.L.V. Pouwels, F. Berger, C. Alix-Panabières, W. Jacot, J.Y. Pierga, F.C. Bidard, M.J. IJzerman

**Author notes:** ***Correspondence:*** Professor Maarten IJzerman University of Melbourne Centre for Cancer Research, School of Population and Global Health.

## Abstract

Patients with metastatic, Estrogen Receptor (ER) positive, HER2-negative, breast cancer, before initiating CDK4/6 inhibitors, receive either single agent endocrine- or chemotherapy based on their clinical risk. In this first-ever trial-based economic evaluation of Circulating Tumor Cells (CTCs), the cost-effectiveness of standardizing the prescription of endocrine- or chemotherapy using a CTC count threshold (with >5 CTCs/7.5mL indicative of unfavorable disease outcomes) was compared to current clinical practice. N=755 ER+ HER2-patients, enrolled in 17 French centres, were randomized to CTC guided or standard of care and were treated according to either through the CTC score or clinical examination. Health state utilities were calculated by mapping the QLQ-C30 to EQ-5D utilities and used to calculate Quality-Adjusted Life Years (QALY) over a 2-year time horizon. Bootstrapping and additional sensitivity analyses were performed to quantify the impact of uncertainty. Health outcomes in both arms were similar, but costs were higher in the CTC guided arm (€19,403) compared to the usual care (€18,254), resulting in an ICER of €104,078/QALY in favor of usual care. However, when the analysis was performed for the clinically high- and low-risk groups separately, CTC enumeration could be a dominant strategy (cost saving) if treatment is de-escalated in clinically high-risk patients as indicated by CTC scores. However, the current analysis was based on the PFS and OS data reported in 2021 and long-term Overall Survival data is collected since then (JCO, 2023 in press). A further analysis of the health economic impact of CTC enumeration in clinically low and high-risk groups is therefore indicated.

## 1. Introduction

For several subtypes of breast cancer that express a particular protein or receptor, targeted therapies have become available which substantially improve survival for these patients (1). However, metastatic breast cancer (MBC) still largely is an incurable condition, with a median survival of only 2 years after detection of metastases (2,3). To extend survival for these patients, it is important to classify the type of tumor and select the optimal treatment as early as possible (4,5). Currently, characterization and staging of breast cancer tumors is done by using histopathological test results from tissue biopsies, imaging modalities or combinations of those. For most subtypes of breast cancer, clinical guidelines already include recommendations for the most effective treatment strategy (6,7).

In metastatic estrogen receptor (ER) positive, HER2-negative breast cancer patients, physicians can prescribe either chemotherapy or endocrine therapy. However, given the palliative setting of MBC, endocrine therapy is the preferred treatment in hormone receptor positive, HER2-negative MBC as confirmed by consensus reports (7,8). A Cochrane systematic review concluded that a policy of treating with endocrine therapy first is recommended, unless there is rapidly progressing disease (9). These exceptions are acknowledged and therapy choice for chemotherapy is predominantly based on the absence of visceral crisis or adverse prognostic factors. Despite these guidelines, and the introduction of CDK4/6 inhibitors, there is significant practice variation and a number of real-world data studies have shown that 15-50% of ER-positive, HER2-negative MBC patients are treated with chemotherapy as a first-line therapy (10,11).

To reduce potentially undesirable practice variation or overtreatment that likely arise due to subjective judgements, standardized objective criteria to guide these treatment decisions should be developed. Liquid biopsies, and circulating tumor cells (CTCs) in particular, have repeatedly shown to have prognostic value for breast cancer patients and may therefore support and improve the standardization of these treatment decisions (12-17). Despite the evidence demonstrating prognostic validity of CTCs, limited studies have actually shown the utility of CTCs to improve outcomes. The SWOG s0500 study was one of the first randomized studies evaluating the use of CTCs to guide a therapy switch after first-line treatment in MBC (18). However, no incremental health benefit could be demonstrated from this study and the main conclusion was that a more effective 2^nd^ line chemotherapy was needed.

The STIC CTC METABREAST trial (STIC trial, NCT01710605) investigated whether guiding endocrine- or chemotherapy by using the number of CTCs as a threshold, is non-inferior to guiding this decision based on patient- and tumor characteristics and the physicians’ expert opinion (19). Recently, the results of the STIC trial were published and demonstrated that CTCs may be a reliable biomarker to guide the use of either chemo- or endocrine therapy in metastatic ER+, HER2-MBC (20). In addition, it was demonstrated that patients that were classified as high risk, either clinically driven or CTC driven (≥5 CTCs per 7.5mL of blood), and receive chemotherapy have a significantly longer progression free survival (PFS) and overall survival (OS) than patients receiving endocrine therapy. Usually, the aim of performing a non-inferiority trial is to demonstrate that especially toxic treatment can be safely de-escalated. The STIC trial demonstrated, however, that the CTC driven strategy may also result in longer PFS when escalating treatment from endocrine- to chemotherapy when ≥5 CTCs were found in patients who were initially clinically identified as low risk.

In addition to the clinical trial, this study aims to investigate the health economic impact of using CTCs as an additional risk-stratifying biomarker in the management or ER+, HER2-MBC. The primary hypothesis is that de-escalation of therapy, from chemotherapy to endocrine therapy as a first-line option, based on an objective measurement would decrease healthcare resource use and reduce chemotherapy related toxicity while not reducing PFS and OS. The analysis of cost-effectiveness is critical to support translation and reimbursement of new technologies deemed effective and empirical cost-effectiveness analysis based on data collected alongside a clinical trial are strongly recommended (21). This study therefore evaluates the health economic impact of standardizing treatment decisions on endocrine and chemotherapy using a CTC count threshold from a healthcare perspective.

## 2. Materials and Methods

### Clinical trial data

Patient-level data were obtained from the randomized controlled STIC trial (20). In this prospective, multi-center trial, patients diagnosed with metastatic disease eligible for their first line of treatment were included and followed for 2 years. For all patients included in the study, the treating physician initially determined whether these patients were eligible for either chemotherapy or endocrine therapy based on the absence of visceral crisis or other adverse prognostic factors. Subsequently, for all patients a CTC test was performed to determine the number of CTCs in 7.5 mL of blood. Following, these patients were randomized over two groups, the (A) Usual Care strategy with physician’s prescribing either single agent endocrine therapy (with no CDK4/6 inhibitor) or chemotherapy based on patient- and tumor characteristics, and (B) the CTC guided strategy with single agent endocrine therapy used in patients with <5 CTCs or chemotherapy when the number of CTCs is ≥5. The number of CTCs found in patients in the usual care arm “A” was not disclosed to the treating physician. In figure 1, an overview of the patients included in each subgroup in both arms is presented.

**Figure 1:**
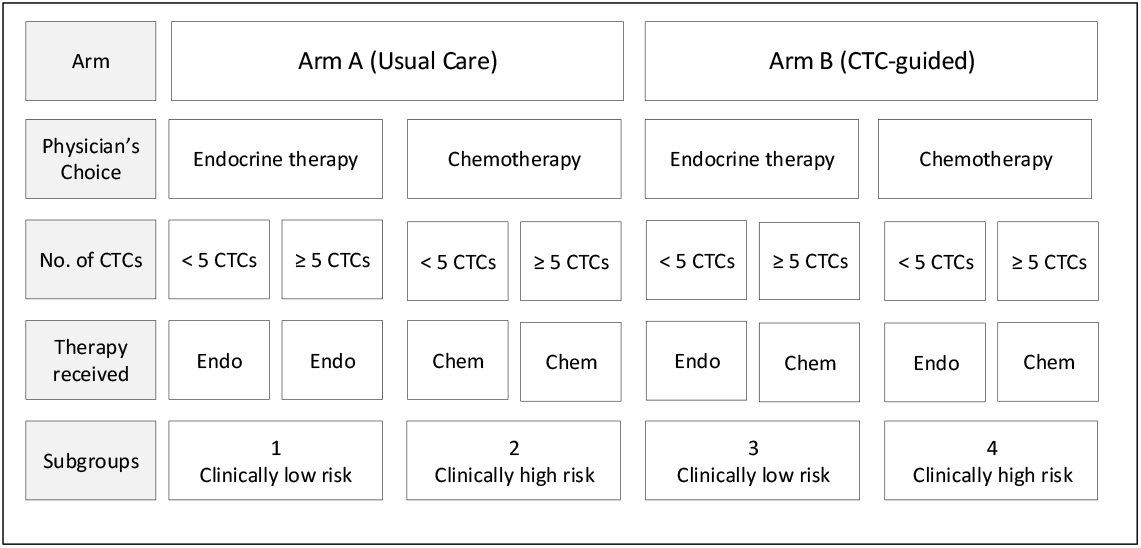
Overview of the treatment allocation decisions of patients included in the STIC Trial Endo = endocrine therapy, Chem = chemotherapy

The main analysis included the per protocol trial population, allowing a head-to-head comparison of the usual care (treatment according to physician choice) and the CTC-guided strategy. Additional subgroup analyses were performed for the clinically low and high-risk subgroups separately. In the clinically low-risk subgroup, the health economic consequences of using CTCs to inform the decision to escalate from endocrine therapy to chemotherapy will be evaluated. In the clinically high-risk subgroup, the health economic consequences of using CTCs to inform the decision to de-escalate from chemotherapy to endocrine therapy will be evaluated.

### Evaluation points

Clinical patient- and tumor characteristics were evaluated at inclusion, every 2 months in the first year and every 4 months in the second year, resulting in a total of 9 time points at which data were collected. In addition, data on the number and type of imaging tests performed, toxic reactions, hospitalization and productivity losses were collected by using a customized evaluation form (designed for the STIC trial) at each of these time points. Data on quality of life were collected using the Quality of Life Questionnaire C30 (QLQ-C30), developed by the European Organization for Research and Treatment of Cancer (EORTC) (22). These questionnaires were completed at inclusion, 2 months, 6 months, 12 months and 24 months.

### Economic evaluation and outcome measures

A cost-utility analysis of using CTCs to standardize the decision to guide either endocrine- or chemotherapy, compared to usual care, was performed. The main outcome measure was the incremental cost-effectiveness ratio (ICER), which is based on the incremental costs divided by the incremental health outcomes in terms of quality adjusted life years (QALYs) of the CTC guided strategy compared with usual care. A willingness-to-pay (WTP) threshold of € 20,000 per QALY was used, indicating that ICERs less than € 20,000 per QALY were considered cost-effective.

The economic evaluation was performed from a healthcare perspective, collecting healthcare resources directly related to the treatment. Cost and outcomes were discounted at a rate of 3%.

#### Health outcomes

The QLQ-C30 incorporates 30 questions classified in 5 functional and 3 symptom scales. As this questionnaire does not allow immediate calculation of health-related quality of life scores (i.e. utility values) a previously published mapping algorithm was used to convert questionnaire answers to utility values (23). When answers on multiple QLQ-C30 questions are missing, the EORTC recommends to impute this data (22). Imputation of missing values was performed in questionnaires with 15 or more questions filled in answers. Multiple imputation was performed in R (version 3.5.1) using the mice package, and was conducted separately for each point in time at which patients should have completed the QLQ-C30 (24). For every time point at which the QLQ-C30 administered, 10 datasets were imputed. In each imputation, all question answers were used as predictors to missing answers. For patients with full answers, and patients for whom these 10 datasets could be imputed, the QLQ-C30 scale scores were mapped to utilities using the response mapping model of Longworth et.al (23)(25). The final utility value at each time point, was defined as the average of the utilities calculated from the 10 imputed datasets. For patients with less than 15 questions answered in a questionnaire, the QLQ-C30 answers were not imputed, and no utility could be calculated from the answers. Therefore, missing utilities were imputed directly for these patients based on the utility derived from the next or previous questionnaire, the arm in which patients were included, whether or not they received chemotherapy, whether any toxicities were experienced in the period before the particular evaluation, progression free survival (PFS) and overall survival (OS) time, the number of CTCs and the age of the patients.

QALYs were calculated by integrating the utility values of patients over the time period in which they experienced these utility values, in a stepwise approach. For example, if a patient was alive for the whole period between the first and the second completion of the QLQ-C30, the average utility over that period was multiplied with the length of that period. If a patient died within a specific period, the average of the utility from the last completed QLQ-C30 and the utility of being dead (utility: 0) was multiplied with the length of the period the patient was still alive after this last completed QLQ-C30.

#### Costs

List prices were derived from the French Healthcare Authority and were used to allocate costs to the imaging tests used and therapies prescribed during each evaluation period. Drug costs for endocrine therapy were calculated by multiplying the list price for a particular drug with the usually prescribed dose of that particular drug in a standard treatment cycle or time period. When no specific treatment switch or stopping date was available, it was assumed that the treatment was continued until progression. Total drug costs were calculated by multiplying the drug costs with the number of cycles the drug was used.

In France, the costs of all hospital stays are registered to diagnosis-treatment-combinations, for which full treatment prices are available independent on the ward in which the patient stayed in the hospital. For those patients in whom the cause of unplanned hospital stays during the evaluation period were unknown, Dutch list prices were used to estimate the cost of the hospital based on the length of stay at the ward in the hospital. Specifically, the Dutch costs of hospital stays were chosen to base these estimations on, as the Dutch healthcare system has a similar diagnosis-treatment-combination coding system, but also provides an overview of the costs of hospital stays in several departments of the hospital. As some of these costs were derived from reference prices from different years, these costs were adjusted to 2019 costs by using Dutch consumer price index levels (CBS, 2015).

Total costs calculated per patient included the cost of treatment, CTC detection (including all materials necessary for the analysis of the results), imaging and hospital monitoring (consultations, medical procedures, possible complications or toxicities and hospitalizations). The exact costs that were allocated to hospital stays, treatments prescribed, imaging techniques used and management of (potential) toxic reactions, are presented in appendix A of the supplement.

### Sensitivity analysis

In the STIC trial, multiple centers in France were included until the final number of patients needed to obtain sufficient statistical power was reached. To reflect the uncertainty in the health and economic outcomes caused by variation on patient-level, a bootstrap analysis, stratified by treatment allocation was performed. In each bootstrap sample, the same number of patients as initially included in each trial arm were randomly sampled with replacement. For each sampled dataset, the incremental costs and QALYs of the CTC-guided arm compared with the usual care arm were calculated. The bootstrap was repeated 10,000 times. Bootstrapping was also performed for the clinically low-risk and the clinically high-risk subgroups. The bootstrap samples were used to calculate the mean health economic outcomes and their corresponding 95% confidence intervals (CI). Cost-effectiveness acceptability curves (CEAC) (26), which determine the probability that the CTC-guided strategy is cost-effective compared to usual care at different willingness-to-pay threshold per QALY, were drawn based on the bootstrap analyses of the full population and both low- and high-risk subgroups. An incremental cost-effectiveness plane (ICEP) was developed to visually demonstrate these results.

The impact on the results of varying prices was investigated through one-way deterministic sensitivity analyses. For this analysis, the prices of hospitalization, and imaging were varied multiple times one-by-one at 10% and 25%. For all analyses, 5,000 bootstrap samples were drawn to estimate mean outcomes. Since varying the costs of each single treatment separately would probably not influence the results, all chemotherapy, hormonal therapy, and targeted therapy prices were varied at once by 10% and 25% in these analyses. The results of the ten most influential cost inputs were demonstrated in tornado diagrams. To perform these analyses, the results of the cost effectiveness analysis were converted to incremental net monetary benefits (iNMB, iNMB = incremental QALY * willingness-to-pay threshold – incremental costs).

## 3. Results

Eligible patients from multiple centers (n=17) in France were included (n=755) and randomized over the usual care arm (n=378) and the CTC guided arm (n=377). The number of patients alive for whom 15 or more questions were unanswered in the QLQ-C30 questionnaires increases over time, with a total number of n=72 (9.54%) for the first questionnaire (timepoint 1), n=186 (24.64%) for the second questionnaire (timepoint 2), n=288 (38.15%) for the third questionnaire (timepoint 3), n=421 (55.76%) for the fourth questionnaire (timepoint 4) and n=571 (75.63%) for the fifth questionnaire (timepoint 5), respectively.

In table 1, the number of patients, mean survival outcomes, QALYs and costs are presented for both trial arms. Guiding chemo- or endocrine therapy based on CTCs results in slightly higher average QALYs (difference of 0.011) and higher costs (difference of € 1,149). These health-economic outcomes result in an ICER of € 104,078 per QALY gained, which exceeds the applied cost-effectiveness threshold of € 20,000 per QALY. Table 1 also presents the 95%-CIs based on the bootstrap sample for both the QALYs and costs.

**Table 1:**
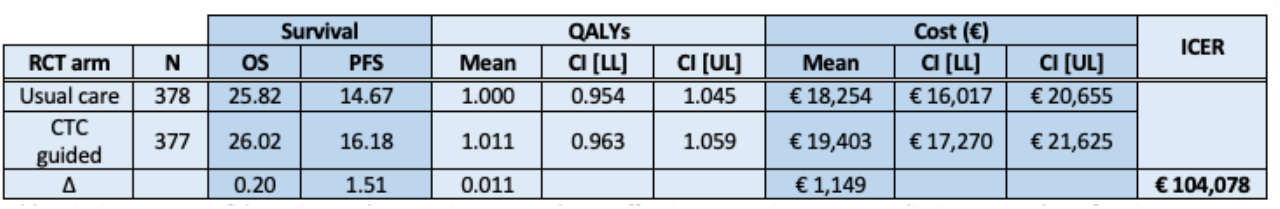
Mean health and economic outcomes with confidence intervals per arm, based on the bootstrap samples. Abbreviations: CI = confidence interval; ICER = incremental cost-effectiveness ratio; LL = Lower limit; N = number of patients; OS = mean overall survival (months); PFS = mean progression-free survival (months); QALY = quality-adjusted life years; UL = Upper limit

Additional results are presented in the supplements including the ICEP for the entire group (figure s1) and the tornado diagrams presenting results if model parameters are changed with either 10% or 25% increase and decrease (figure S2a and b). The sensitivity analysis shows the results are robust, even if parameter values are changed with 25%. Chemotherapy prices are the main driver in the economic model. Table 2 presents all the results for the clinically high- and low-risk subgroups (defined in figure 1). This includes the number of patients, the mean survival outcomes, QALYs and costs.

**Table 2:**
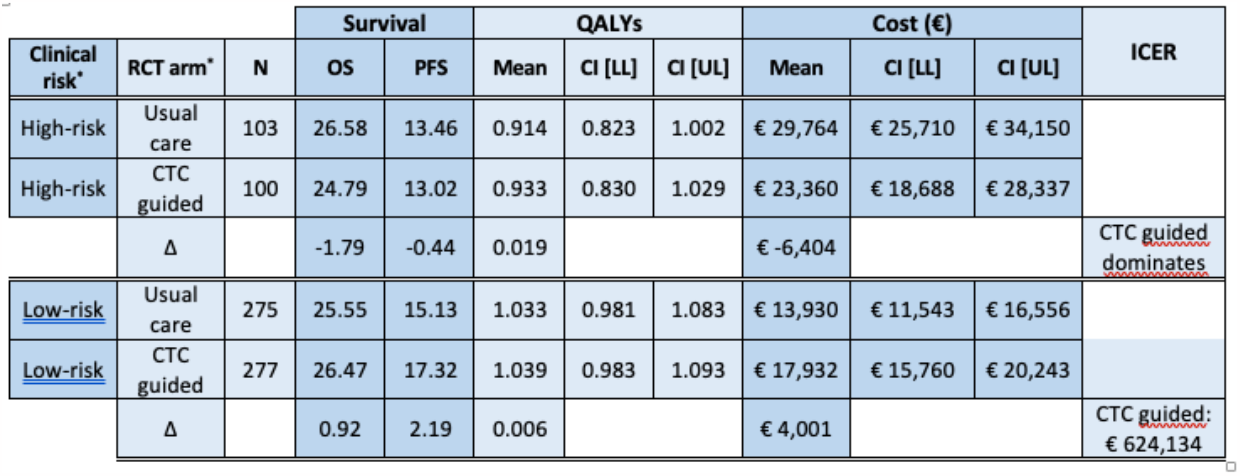
Mean health and economic outcomes with confidence intervals for the clinically high- and low-risk subgroup based on the bootstrap samples. Abbreviations: CI = confidence interval; ICER = incremental cost-effectiveness ratio; OS = mean overall survival (months); PFS = mean progression-free survival (months); QALY = quality-adjusted life years; UL and LL = upper and lower limit. * Subgroups clinical risk were defined in figure 1.

In the clinically low-risk subgroup, the median PFS is 17.2 months (95%CI, 15.5 – 20.2) in the CTC guided arm compared to 15.3 months (95%CI, 11.7 – 17.2) for the physician choice arm. The QALY difference between these groups is 0.006 (1.039 CTC guided arm versus 1.033 in the physician choice arm) and the cost difference is € 4,001, resulting in an ICER of € 624,134 per QALY gained (table 2, clinically low-risk patients).

For the high-risk subgroup, patients initially selected by the physician for chemotherapy can be de-escalated to endocrine therapy if less than 5 CTCs were detected. The clinical results in the trial demonstrate that median PFS is not different for high-risk patients treated with chemotherapy according to the physician choice (12.8 (95%CI, 10.8 – 15.4)) compared to high-risk patients where CTCs were used to select either chemo- or endocrine therapy (11.2 (95%CI, 8.6 – 15.3). Table 2 also presents the QALY difference between these two groups, which is 0.019 (0.933 in the CTC guided arm vs. 0.914 in the physician choice arm). The cost difference between these groups is € -6,404, which eventually leads to a dominant strategy in favor of the CTC guided strategy.

### Deterministic sensitivity analysis

To assess the uncertainty surrounding the calculated ICER, bootstrapping with 10,000 replications was performed. The incremental cost-effectiveness plane for each subgroup is presented in figure 2, demonstrating the cost differences between the risk-groups to be the main difference, and the opportunity for CTC guided arm to be cost-saving without impacting health outcomes in the clinically high-risk subgroup. The results of the sensitivity analysis for the entire population and the clinically low- and high-risk groups are presented in the supplement (figure s2ab, s3ab, s4ab). These results demonstrate the outcome to be robust to changes in prices.

**Figure 2:**
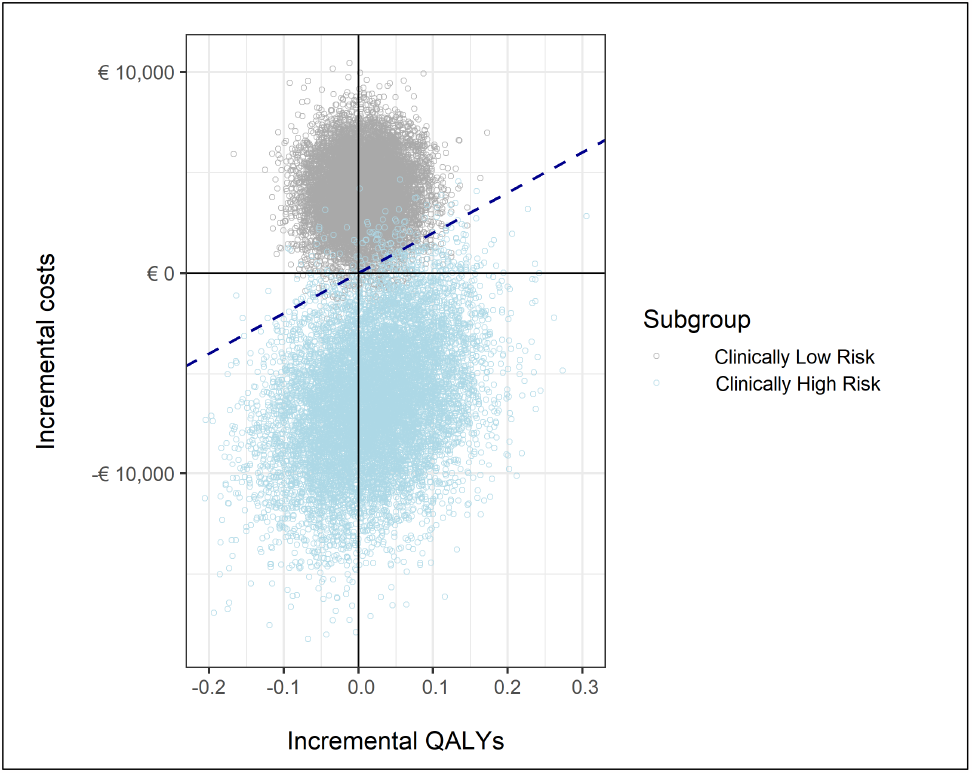
Incremental costs and QALYs based on the bootstrap samples in the clinically low- (grey) and high-risk (light blue) population (n=10,000) indicating potential cost-savings using CTC enumeration in the clinically high-risk group.

### Cost-effectiveness acceptability (CEAC)

The CEACs that were created based on bootstrap sampling are presented in figure 3. The CEAC depicts the probability of a strategy being cost-effective at different WTP thresholds. In the base-case, a WTP of €20,000 per QALY is assumed which results in the probability of CTCs being cost-effective of 29.04%. If the WTP is increased, the probability of CTCs being a cost-effective strategy increases to 39.46% (WTP=€50,000/QALY) and 46.25% (WTP=€80,000/QALY).

**Figure 3:**
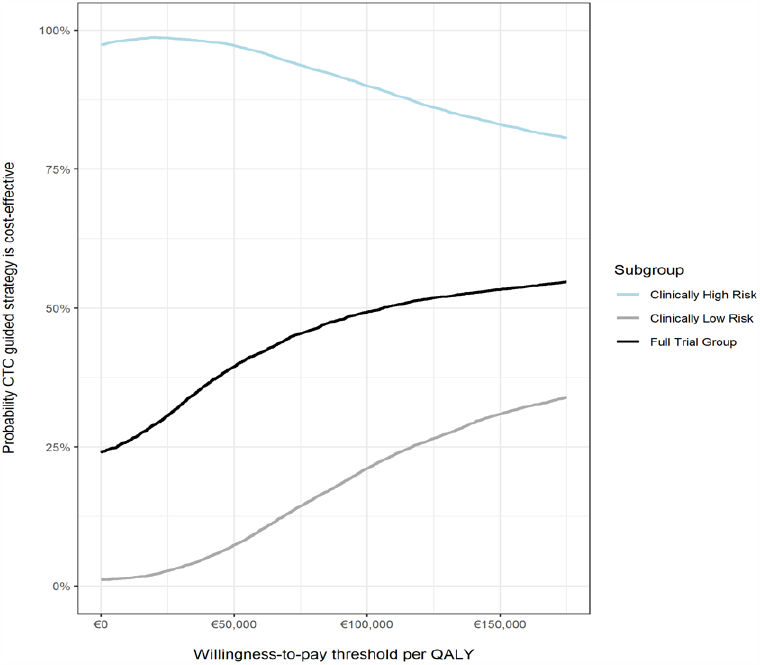
Cost-Effectiveness Acceptability Curves for the full trial group, and for the clinically low- and high-risk subgroups.

More interesting is the evaluation of the probability of being cost-effective if the use of CTC guidance is restricted to the clinically high-risk population. In that case, the probability of being cost-effective is 98.74% if a WTP of € 20,000/QALY is used, a figure that is slightly less (92.97%) for a WTP of € 80,000/QALY. For the low-risk patients, the probability CTC guidance will be a cost-effective strategy is only 15.7% at a WTP of € 80,000/QALY.

## 4. Discussion

Although the STIC trial demonstrated non-inferiority of using the CTC-guided strategy to provide more standardized guidance in the selection of either single agent endocrine- or chemotherapy, this study further explores the health economic impact. Of note, the recent use of CDK4/6 inhibitor in combination with endocrine therapy has considerably changed both the efficacy and the cost of first line therapy. Nevertheless, if the entire population is considered, the analysis showed that incremental QALYs of using CTC guided vs. standard of care, ranges from -0.2 to 0.2, while the incremental costs range from € -20,000 to € 10,000. Consequently, this cost-utility analysis of the entire patient population resulted in an ICER of € 104,000/QALY, indicating that using CTCs to guide chemo- or endocrine therapy is not cost-effective.

However, as the STIC trial is designed to be a non-inferiority study with only marginal clinical benefits, the ICER is relatively uninformative because of the negligible clinical difference. One explanation is that the follow-up of the STIC trial, censored to 2 years, may have been too short to generate a positive outcome in terms of the estimated incremental ICER gain. For instance, even though there is a disutility of escalating from endocrine therapy to chemotherapy when ≥5 CTCs were detected, this difference may not outweigh the potential long-term survival benefits. Likewise, the utility gain derived when de-escalating treatment from chemotherapy to endocrine therapy when <5 CTCs were detected, may not outweigh the additional costs of using CTCs to guide this treatment. Although trial outcomes beyond this 2 year time horizon could be extrapolated using statistical modelling techniques, collecting data on the actual long term survival and quality of life of patients would be preferred yet will take some years.

In addition to the analysis of the entire population, it may also be clinically relevant to consider the subgroups of clinically high and low risk. From a clinical perspective, new diagnostic tests are more likely to be used to guide the escalation of treatment in clinically low-risk patients than for the de-escalation of therapy in clinically high-risk patients, mainly due to concerns of undertreatment. It therefore is clinically relevant to compare both subgroups, while appreciating the study was not designed neither powered to look into these subgroups.

The clinical benefits of treatment escalation in the low-risk group with ≥5 CTCs detected is relatively small with 0.006 QALYs gained. Therefore, the health economic analysis failed to demonstrate a clear benefit with ICERs up to €624,134/QALY.

In contrast, however, from a health economic perspective it was clearly demonstrated that de-escalating treatment from chemo- to endocrine therapy in the high-risk group is a dominant strategy, implying it is a cost-saving strategy with better clinical outcomes. However, it is less likely that such treatment de-escalation will be adopted in clinical practice, mainly because of concerns of under-treatment.

Demonstrating cost-effectiveness is widely considered a critical component in the translation and implementation of new treatments and diagnostic strategies. Several economic evaluations of biomarker tests for breast cancer were published, all related to risk-stratification and decisions to start systemic treatments (27-29). However, only one study is based on prospective randomized clinical trials. To our knowledge, the current study is the first trial-based economic evaluation investigating liquid biopsies. While RCTs are the preferred study design, one of the main challenges is the time lag between the design and analysis of the results and, hence, the change in treatment options and clinical guidelines since the start of the trial. While the actual clinical data may therefore be less informative, the economic model and its underlying principles are informative for future studies.

Although the STIC trial was designed as a clinical trial allowing an economic evaluation, several statistical methods were used to estimate the health economic outcomes. The QLQ-C30 questionnaire was mapped to utilities using the algorithm of Longworth et. al. (23). Obviously, direct measurement of EQ5D or using a different algorithm could change the estimated QALY gain and subsequently the health economic outcomes. Furthermore, in the analysis of the full trial population, list prices were used to calculate patient-level costs. As those list prices may vary substantially over time and over different countries, these may not be the best estimate of the actual treatment costs. Therefore, this uncertainty was addressed in the sensitivity analysis demonstrating a significant impact of chemotherapy followed by targeted therapy prices.

Finally, in the STIC trial, a CTC count threshold of 5 CTCs per 7.5 mL blood was used to either prescribe chemotherapy (≥ 5CTCs) or endocrine therapy (<5 CTCs) in the CTC guided strategy. The CTC count of 5 is based on previously developed clinical prognostic models [12]. As this cut-off point can also influence the health economic outcomes, future work may elaborate on the impact of different CTC thresholds.

## 5. Conclusions

Using CTC counts to guide endocrine and chemotherapy currently appears unlikely to be cost-effective on the short term when used in all ER+ HER2-MBC patients. However, using CTCs to de-escalate from chemotherapy to single agent endocrine therapy in the clinically high-risk subgroup could be cost-saving without affecting health outcomes. The bootstrap analysis showed this can be a dominant strategy, with 98% probability to be cost-effective assuming a willingness to pay of €20,000/QALY.

The STIC trial also demonstrated that escalating treatment from single agent endocrine to chemotherapy in the clinically low-risk subgroup appeared to be beneficial in terms of clinical outcomes. However, from a health economic perspective it is unsure that this strategy will be cost-effective (€ 624,134/QALY) in the short term, mainly because increased use of chemotherapy will be more costly. Only recently, 2023 data on Overall Survival are collected by the lead investigators of the STIC trial (Bidard et al, JCO, 2023 in press) and thus the results of this health economic study based on the 2021 data need to be updated accordingly.

## Data Availability

All data produced in the present work are contained in the manuscript

