## Supplemental files for "Cost-effectiveness of CTC guided chemo- or endocrine therapy in ER+ HER2- metastatic breast cancer – results from a randomized controlled multicenter trial"

### Appendix A. Allocation of costs and sources

| Type of costs | Costs accounted | Unit | Source |
|---|---|---|---|
| Alkeran | € 402.67 | Cycle | French Healthcare Authority (list prices) |
| Capecitabin | € 402.67 | Cycle | |
| Carboplatin | € 402.67 | Cycle | |
| Cyclophosphamide | € 402.67 | Cycle | |
| Docetaxel | € 402.67 | Cycle | |
| Doxorubicin | € 402.67 | Cycle | |
| Doxo liposomal | € 402.67 + € 1320.90 | Cycle | |
| Epirubicin | € 402.67 | Cycle | |
| Eribulin | € 402.67 + € 1504 .00 | Cycle | |
| Fluorouracil | € 402.67 | Cycle | |
| Methothrexate | € 402.67 | Cycle | |
| Paclitaxel | € 402.67 | Cycle | |
| Vinorelbin | € 402.67 | Cycle | |
| Other chemo | € 402.67 | Cycle | |
| Consult | € 28.00 | One appointment | |
| Tamoxifen | € 3.34 | Month | |
| Anti Aromastase :Letrozole | € 41.71 | Month | |
| LH-RH antagonist: Zoladex implant | € 109.49 | Month | |
| Fulvestrant | € 926.68 | Month | |
| Bevacizumab | € 402.67 + € 2130.00 | Cycle | |
| Everolimus | € 2400.47 | Cycle | |
| Aranesp | € 211.00 | Cycle | |
| Eprex | € 143.78 | Cycle | |
| Neorecormon | € 820.60 | Cycle | |
| Lenograstim | € 419.80 | Cycle | |
| Filgrastim | € 560.10 | Cycle | |
| Pegfilgrastim | € 596.92 | Cycle | |
| Intensive care department | € 2,452.18 | Day | Dutch Healthcare Authority (list prices) |
| Conventional hospital department | € 1,314.52 | Day | |
| Daycare Hospital | € 306.66 | Day | |
| Home treatment | € 124.11 | Day | |
| Home care | € 75.50 | Day | |
| X-ray | € 21.28 | Scan | French Healthcare Authority (list prices) |
| Echo | € 52.45 | Scan | |
| Bone scan | € 168.71 | Scan | |
| PET scan | € 1,089.54 | Scan | |
| CT scan | € 183.54 | Scan | |
| MRI | € 240.09 | Scan | |
| Radiotherapy | € 4,386.44 | Full treatment | |
| Cost CTC Test | € 500.00 | Test | Based on micro-costing in one of the participating centres. |

**Appendix B Incremental cost-effectiveness plane for the entire trial population**

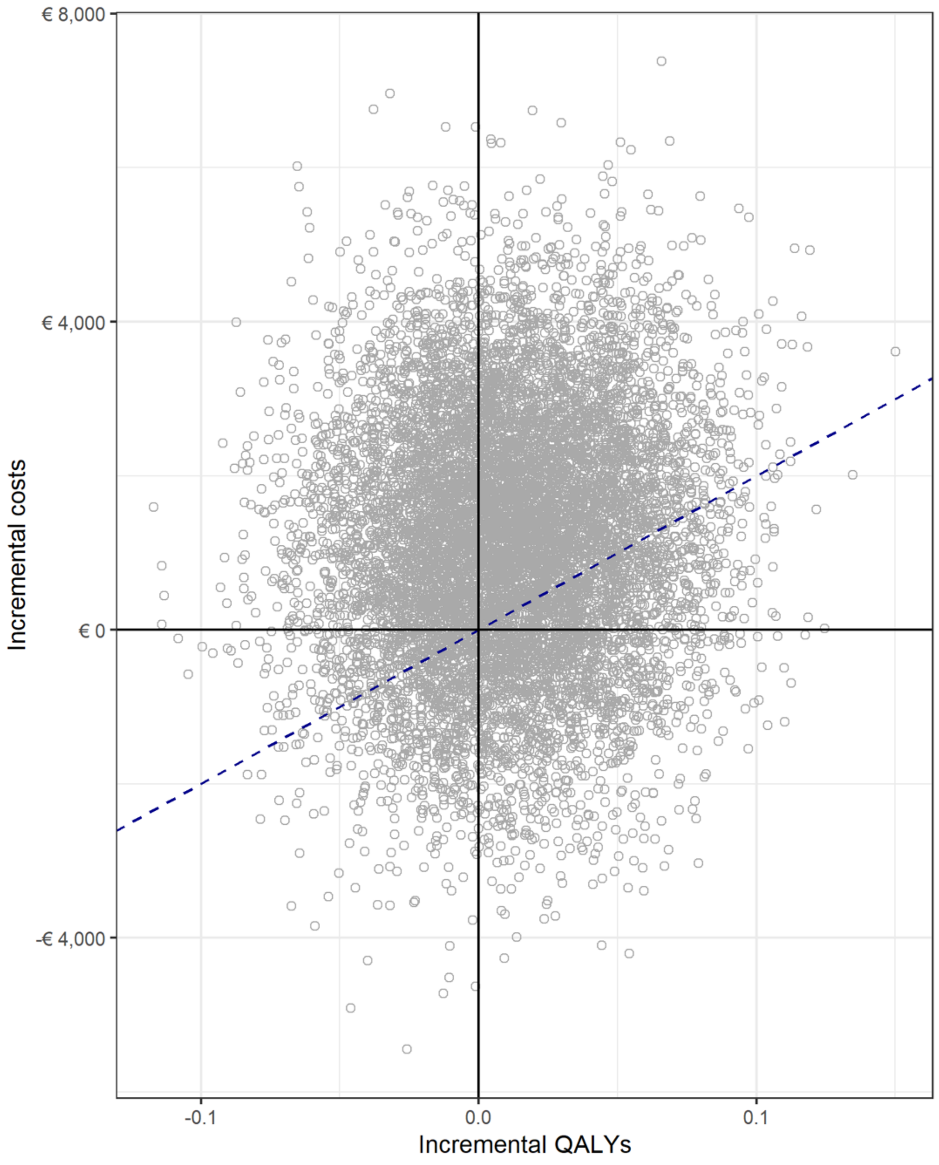


**Figure S1.** Cost-effectiveness plane for the entire clinical trial population. Summary data is presented in table 1 in the manuscript, with an incremental cost-effectiveness ratio (ICER) of €104,078/QALY.

**Appendix C: Sensitivity analyses**

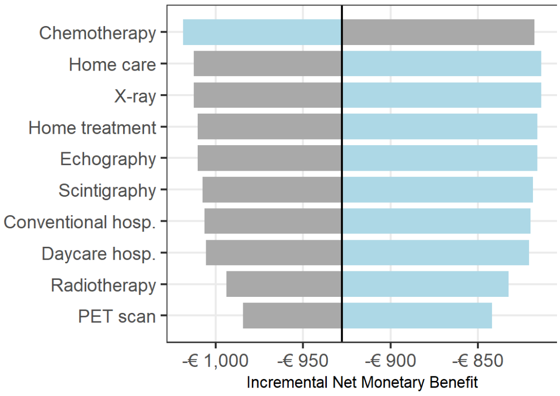


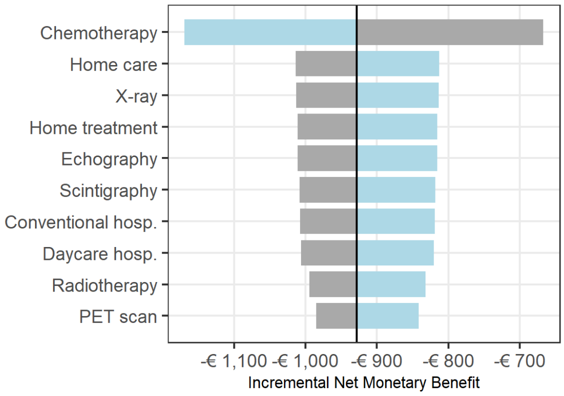


**Figure S2a and b.** Sensitivity analysis on the main cost components influencing the outcome for the *entire trial population*. The cost inputs are changed with 10% (left) and 25% (right) respectively. Results are presented in Net Monetary Benefits, calculated assuming a WTP of € 20,000/QALY. Negative values mean an incremental cost per QALY relative to the WTP.

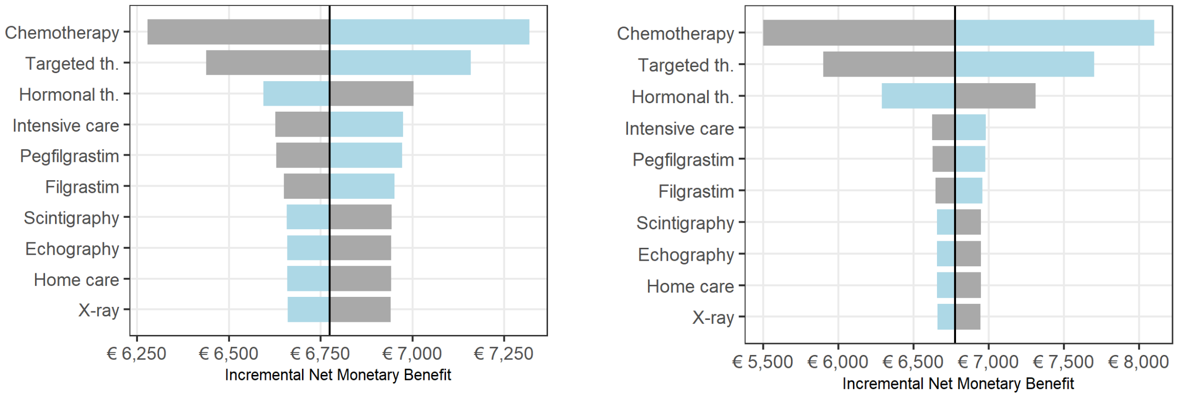


**Figure S3a and b.** Sensitivity analysis on the main cost components influencing the outcome for the clinically *high-risk* population. The cost inputs are changed with 10% (left) and 25% (right) respectively. Results are presented in Net Monetary Benefits, calculated assuming a WTP of € 20,000/QALY. Positive values mean a cost saving per QALY relative to the WTP. This implies the results are robust to changes in cost of model inputs.

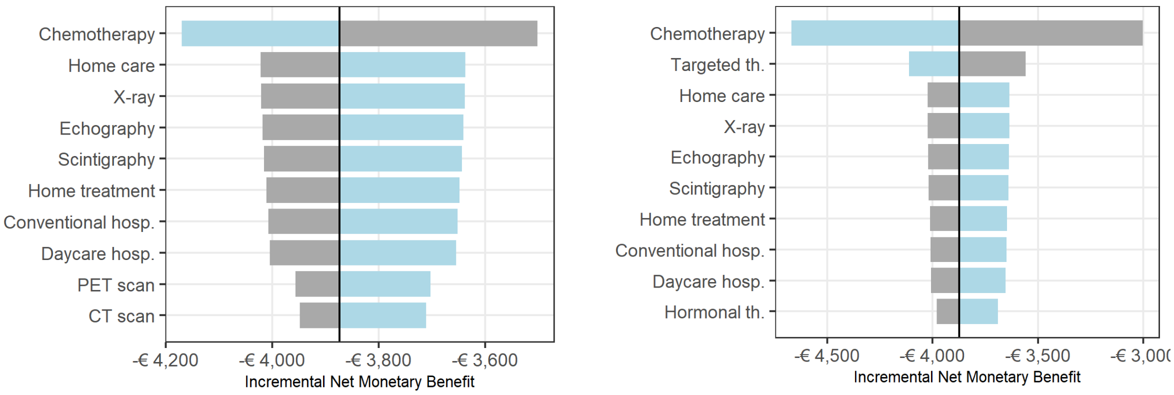


**Figure S4a and b.** Sensitivity analysis on the main cost components influencing the outcome for the clinically *low-risk* population. The cost inputs are changed with 10% (left) and 25% (right) respectively. Results are presented in Net Monetary Benefits, calculated assuming a WTP of € 20,000/QALY. Negative values mean a incremental cost per QALY relative to the WTP. This analysis demonstrates the results are robust to changes in cost of model inputs.
